# COVID-19 and associations with frailty and multimorbidity: a prospective analysis of UK Biobank participants

**DOI:** 10.1101/2020.06.09.20126292

**Authors:** S J Woolford, S D’Angelo, E M Curtis, C M Parsons, K A Ward, E M Dennison, H P Patel, C Cooper, N C Harvey

## Abstract

**Background:** Frailty and multimorbidity have been suggested as risk factors for severe COVID-19 disease.

**Aims:** We investigated whether frailty and multimorbidity were associated with risk of hospitalisation with COVID-19 in the UK Biobank.

**Methods:** 502,640 participants aged 40-69 years at baseline (54-79 years at COVID-19 testing) were recruited across UK 2006-10. A modified assessment of frailty using Fried’s classification was generated from baseline data. COVID-19 test results (England) were available 16/03/2020-01/06/2020, mostly taken in hospital settings. Logistic regression was used to discern associations between frailty, multimorbidity and COVID-19 diagnoses, adjusting for sex, age, BMI, ethnicity, education, smoking and number of comorbidity groupings, comparing COVID-19 positive, COVID-19 negative and non-tested groups.

**Results:** 4,510 participants were tested for COVID-19 (positive=1,326, negative=3,184). 497,996 participants were not tested. Compared to the non-tested group, after adjustment, COVID-19 positive participants were more likely to be frail (OR=1.3 [95% CI=1.1, 1.7]), report slow walking speed (OR=1.3 [1.1, 1.6]), report two or more falls in the past year (OR=1.3 [1.0, 1.5]) and be multimorbid (≥4 comorbidity groupings vs 0-1: OR=1.9 [1.5, 2.3]). However, similar strength of associations were apparent when comparing COVID-19 negative and non-tested groups. Furthermore, frailty and multimorbidity were not associated with COVID-19 diagnoses, when comparing COIVD-19 positive and COVID-19 negative participants.

**Discussion and conclusions:** Frailty and multimorbidity do not appear to aid risk stratification, in terms of a positive versus negative results of COVID-19 testing. Investigation of the prognostic value of these markers for adverse clinical sequelae following COVID-19 disease is urgently needed.

## Background

The first case of 2019 Novel Coronavirus (COVID-19) disease (caused by the Severe Acute Respiratory Syndrome Coronavirus 2 [SARS-CoV-2]) was reported in Wuhan, China in December 2019 [1]. Since then, the number of global cases have increased rapidly, with the WHO declaring COVID-19 a pandemic in March 2020 [2]. At the time of manuscript preparation, more than 5 million cases have been confirmed across 213 countries and territories, with more than 300,000 associated deaths [3].

The identification of risk factors for contracting COVID-19 is crucial, in order to inform public health policy and facilitate the appropriate distribution of healthcare resources. Preliminary data from Asia, Europe and the United States suggest that the majority of individuals with COVID-19 are aged >50 years, with most deaths occurring in those aged >60 years [4-7]. Multimorbidity has also been associated with COVID-19 disease, the need for ventilatory support and higher rates of mortality [4-7]. As a result, COVID-19 patients have frequently been described within the academic discourse as “frail”, with this term being used in its more colloquial sense [8-10]. Furthermore, clinical management guidelines, such as those produced by the UK National Institute for Health and Care Excellence (NICE), typically recommend the assessment of frailty as the initial step when triaging suspected COVID-19 patients [11]. However, the clinical syndrome of frailty has yet to be formally examined in relation to COVID-19 disease, beyond single case reports [12].

Therefore, we aimed to examine the associations between COVID-19 diagnoses, frailty and multimorbidity within the UK Biobank, a large prospective community cohort of over half a million UK residents.

## Methods

### Study population

This study was a prospective community-based cohort analysis of UK Biobank participants. Between 2006 and 2010 potential participants were invited for recruitment, with inclusion criteria being registered with a general practitioner, living within reasonable travelling distance of an assessment centre and being aged 40-69 years. 502,640 participants were recruited across 22 assessment centres in England, Scotland and Wales, with a response rate of 5%.

### Baseline characteristics

Information on lifestyle, social history and medical history was collected using a series of computer-based touchscreen questionnaires, followed by face-to-face interviews with trained research staff. Recorded data included sex, age, ethnicity, level of educational attainment, alcohol consumption (never or special occasions only; one to three times per month; one to four times per week; or daily or almost daily), smoking status (never smoked; ex-smoker; or current smoker) and number of falls in the past year (no falls; only one fall; or more than one fall). Height and weight were also measured by trained research staff using a standardised technique, and BMI was subsequently calculated (kg/m^2^).

### Assessment of frailty

We calculated frailty using a modified version of five frailty phenotype indicators originally reported by Fried and colleagues [13], and later adapted for use in the UK Biobank data set [14]. Table 1 outlines the criteria for the frailty indicators used, compared to Fried and colleagues original criteria. The criteria for the low physical activity frailty indicator was further adapted for use in this study, based on a comparable exercise metric within the data set available to us. Grip strength was measured using a Jamar J00105 hydraulic hand dynamometer, with both right and left hands being assessed and the lower result used in the analysis. The other frailty indicators were assessed via self-reported touchscreen questionnaire answers. The associations with multiple falls in the past year with COVID-19 diagnoses was also examined, due to the associations of frequent falls with frailty and the frequency of which a fall is the first presentation of the frailty syndrome to healthcare providers [15,16].

**Table 1.** Frailty indicators originally adapted for use in the UK Biobank by Hanlon and colleagues, based on Fried and colleagues original frailty phenotype.

|  | Cardiovascular Health Study frailty indicators (Fried and colleagues) [13] | Adapted UK Biobank frailty indicators (Hanlon and colleagues) [14] |
| --- | --- | --- |
| Weight loss | Self-reported: "In the last year, have you lost more than 10 pounds unintentionally?"<br>Yes=1, no=0 | Self-reported: "Compared with one year ago, has your weight changed?"<br>Yes, lost weight=1, other=0 |
| Exhaustion | Self-reported: "How often in the last week (a) did you feel that everything was an effort, or (b) could you not get going?"<br>Moderate amount of the time [3–4 days] or most of the time=1, other=0 | Self-reported: "Over the past two weeks, how often have you felt tired or had little energy?"<br>More than half the days or nearly every day=1, other=0 |
| Low physical activity | Self-reported: Minnesota Leisure Time Activity Questionnaire and Kcal of activity per week subsequently estimated.<br>Lowest 20% of cohort=1, other=0 | Self-reported: "In a typical week, on how many days did you do 10 minutes or more of moderate physical activities like carrying light loads, cycling at normal pace? (do not include walking)" *<br>0-1 day/week=1, other=0 |
| Slow walking speed | Measured time to walk 15 feet.<br>Lowest 20% of cohort=1, other=0 | Self-reported: "How would you describe your usual walking pace?"<br>Slow=1, other=0 |
| Grip strength | Measured grip strength, adjusted for sex and body-mass index.<br>Lowest 20% of cohort=1, other=0 | Measured grip strength.<br>Sex and body-mass index adjusted cut-offs taken from Fried and colleagues. |

\* Criteria not from Hanlon and colleagues, and based on comparable data available for use for the purposes of this study.

As per Fried and colleagues, participants were classified as not frail (0 frailty indicators), pre-frail (1-2 frailty indicators) or frail (≥3 more frailty indicators). Frailty status was not calculated for participants with missing data for three or more frailty indicators, as per Fried and colleague’s methodology.

### Assessment of multimorbidity

Participants reported doctor-diagnosed chronic health conditions during face-to-face interviews at study recruitment, apart from cancer, which was reported during touchscreen questionnaires. To avoid repeated counting of closely related or clinically similar chronic health conditions, comorbidities have been categorised according to 43 comorbidity groupings, originally established for a large epidemiological study in Scotland [17], and subsequently amended for use in the UK Biobank [18]. Supplementary Table 1 shows the full list of comorbidity groupings and the corresponding health conditions. Number of comorbidity groupings were then summed, and categorised (0-1; 2; 3 or ≥4 comorbidity groupings).

### COVID-19 testing

COVID-19 diagnoses were sourced via available COVID-19 test results within the UK Biobank data set at the time of manuscript preparation (from 16^th^ March 2020 to 1^st^ June 2020), sourced from Public Health England [19]. The vast majority of these COVID-19 tests were via a combined nasal and throat swab. In intensive care settings, lower respiratory secretion samples were also subject to COVID-19 testing. Samples were transported in a medium suitable for viruses (typically a balanced salt solution), and PCR-based testing was performed.

The time period from which COVID-19 testing data were available for analysis from the UK Biobank was during the peak of the UK COVID-19 outbreak, when the overwhelming majority of COVID-19 testing took place in hospital settings. Therefore, it can be assumed that all those who were tested for COVID-19 presented with symptoms, due to COVID-19 or otherwise, severe enough to warrant hospital admission. The background population group represent those not tested for COVID-19, including those who had did not have COVID-19, as well as undiagnosed COVID-19 cases who were asymptomatic or only had mild symptoms. Current prevalence estimates of undiagnosed COVID-19 cases within the UK community population are approximately 0.3% [20].

### Statistical analysis

Participants were divided into three groups for comparison: 1) participants who tested positive for COVID-19 (COVID-19 +ve group), 2) participants who tested negative for COVID-19 (COVID-19 -ve group), and 3) participants who had not been tested for COVID-19 (background population group). Baseline characteristics of these three groups were analysed by reporting mean (standard deviation, SD) or median (interquartile range, IQR) as appropriate for continuous variables, and number (percentages) for categorical variables. Differences between groups were tested with unpaired t-tests, Mann-Whitney U tests or Pearson Chi-square tests, as appropriate.

Logistic regression was used to explore the associations between COVID-19 +ve vs COVID-19 -ve groups and frailty status, frailty indicators, number of falls in the past year and number of comorbidity groupings, with groups being stratified for age (<60 or ≥60 years at baseline). Covariates considered included sex, age, BMI, ethnicity, educational attainment, smoking status and number of comorbidity groupings (where comorbidity was not the exposure). Similar analyses for COVID-19 +ve and COVID-19 -ve groups vs the background population group were also performed.

All analyses were performed with Stata v 15.1 (StataCorp, College Station, Texas, USA). All UK Biobank participants gave written informed consent for data collection, analysis, and linkage at study recruitment. This study had ethics approval as part of overall UK Biobank ethics approval (NHS National Research Ethics Service 16/NW/0274). We undertook the study under UK Biobank Access Application 3593.

## Results

### Study population

A total of 4,510 UK Biobank participants were tested for COVID-19. Of these, 1,326 tested positive and 3,184 tested negative. 497,996 participants were not tested. 1,769 participants had missing data for three or more frailty indicators, and were excluded from any analyses requiring these data.

### Baseline characteristics

Table 2 shows the baseline characteristics of the three comparative groups. The median age ranged from 58-60 years by group, consistent with the recruitment criteria of ages 40-69 years. Median age at the time of COVID-19 testing was 70 and 71 years, for those who tested positive and negative respectively. The COVID-19 +ve group were more likely to be male, of greater BMI, of non-white ethnicity and lower educational attainment, and less likely to consume alcohol and to have never smoked, when compared with the background population group. They were also more likely to be frail, exhibit a number of frailty indicators (weight loss, exhaustion, slow walking speed and weakness of grip), to report two or more falls in the past year and to report a higher number of comorbidity groupings, when compared with the background population group. However, comparison of COVID-19 -ve group with the background population group yielded a similar pattern of results, apart from being less likely to be male.

**Table 2.**
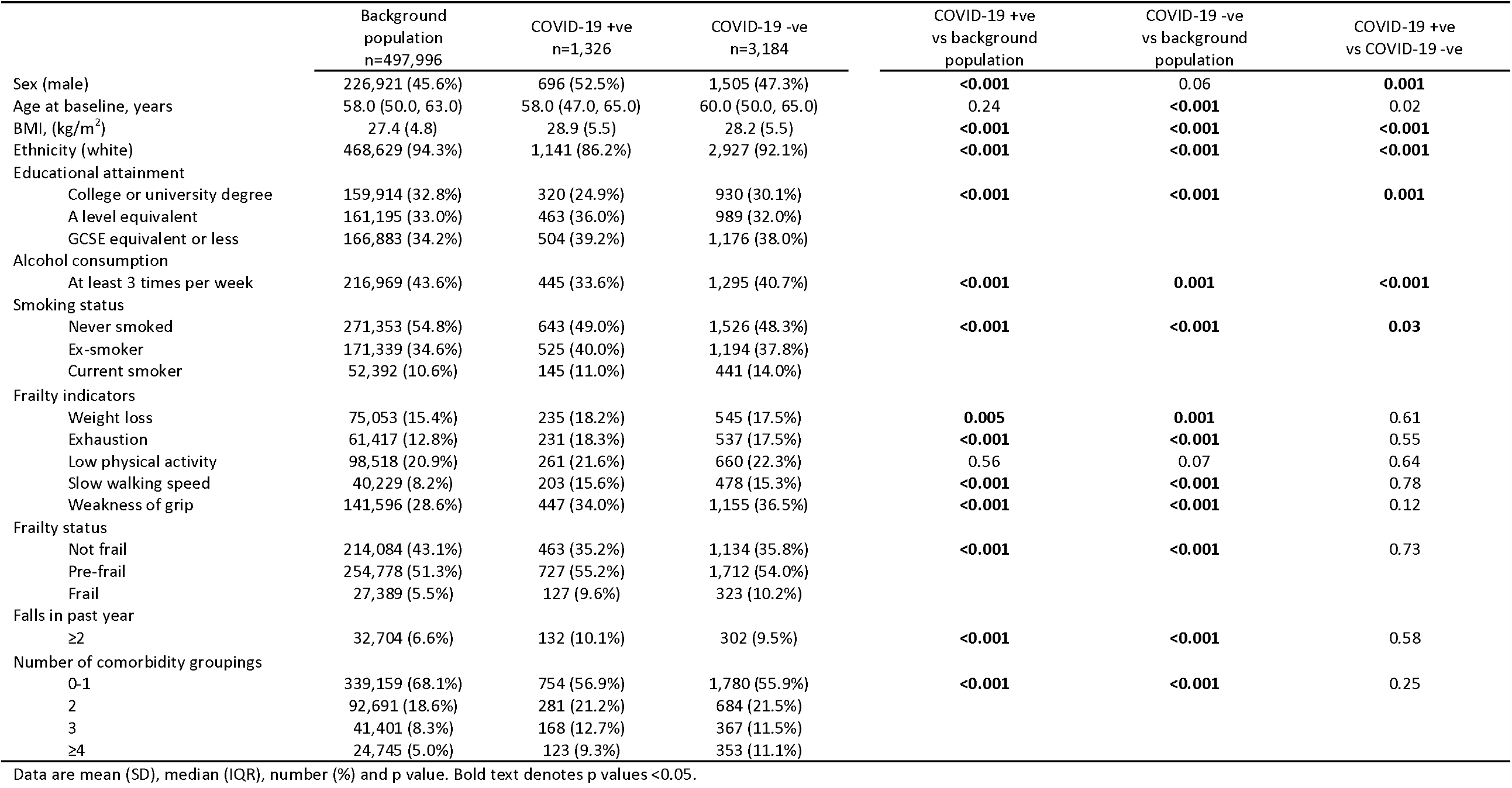
Characteristics of background population, COVID-19 +ve and COVID-19 -ve groups.

When comparing COVID-19 +ve and -ve groups, COVID-19 +ve participants were more likely to be male, of greater BMI, of non-white ethnicity, greater BMI, of lower educational attainment and consume less alcohol than COVID-19 -ve participants. However, the two groups did not differ in terms of frailty status, frailty indicators, falls in the past year or number of comorbidity groupings.

### Associations between frailty, falls, multimorbidity and COVID-19 diagnoses

Table 3 documents the associations between potential COVID-19 risk factors in the COVID-19 +ve group vs the COVID-19 -ve groups, also stratified by age (<60 or ≥60 years at baseline, corresponding to <70-74 and ≥70-74 years at COVID-19 testing). After adjustment for sex, age, BMI, ethnicity, educational attainment, smoking status and number of comorbidity groupings there were no associations between frailty, indicators of frailty, falls in the past year or number of comorbidity groupings and testing positive for COVID-19. Age stratification revealed no additional associations at older or younger ages. However, odds ratios for frailty indicators and multimorbidity were, in general, more likely to be greater than unity in the older age group, compared with those for the younger age group.

**Table 3.** Associations for COVID-19 risk factors in COVID-19 +ve vs COVID-19 -ve groups.

|  | Unadjusted |  |  | Adjusted<sup>1</sup> |  |  |
| --- | --- | --- | --- | --- | --- | --- |
|  | All | <60 years at baseline | ≥60 years at baseline | All | <60 years at baseline | ≥60 years at baseline |
| Frailty status |  |  |  |  |  |  |
| Not frail | Reference | Reference | Reference | Reference | Reference | Reference |
| Pre-frail | 1.1 (0.9, 1.3) | 1.0 (0.8, 1.2) | 1.1 (0.9, 1.4) | 1.0 (0.8, 1.2) | 0.9 (0.7, 1.1) | 1.1 (0.9, 1.4) |
| Frail | 1.0 (0.8, 1.3) | 0.8 (0.6, 1.2) | 1.2 (0.8, 1.6) | 0.9 (0.6, 1.2) | 0.7 (0.5, 1.0) | 1.0 (0.7, 1.5) |
| Frailty indicators |  |  |  |  |  |  |
| Weight loss | 1.1 (0.9, 1.3) | 1.0 (0.8, 1.3) | 1.1 (0.9, 1.4) | 1.1 (0.9, 1.3) | 1.0 (0.8, 1.3) | 1.1 (0.8, 1.4) |
| Exhaustion | 1.0 (0.8, 1.2) | 1.0 (0.8, 1.3) | 1.1 (0.8, 1.4) | 1.0 (0.8, 1.2) | 1.0 (0.8, 1.3) | 1.0 (0.7, 1.3) |
| Low physical activity | 1.0 (0.8, 1.2) | 0.8 (0.6, 1.0) | 1.2 (0.9, 1.5) | 0.9 (0.8, 1.1) | 0.8 (0.6, 1.0) | 1.2 (0.9, 1.5) |
| Slow walking speed | 1.1 (0.9, 1.4) | 0.8 (0.6, 1.1) | 1.2 (1.0, 1.6) | 1.0 (0.8, 1.2) | 0.7 (0.5, 1.0) | 1.2 (0.9, 1.5) |
| Weakness of grip | 0.9 (0.8, 1.1) | 0.9 (0.8, 1.2) | 0.9 (0.7, 1.1) | 0.9 (0.7, 1.0) | 0.9 (0.7, 1.1) | 0.9 (0.7, 1.1) |
| Falls in past year |  |  |  |  |  |  |
| 0-1 | Reference | Reference | Reference | Reference | Reference | Reference |
| ≥2 | 1.1 (0.9, 1.4) | 0.9 (0.8, 1.1) | 1.0 (0.9, 1.1) | 1.1 (0.9, 1.5) | 0.9 (0.8, 1.1) | 1.0 (0.9, 1.2) |
| Number of comorbidity groupings |  |  |  |  |  |  |
| 0-1 | Reference | Reference | Reference | Reference | Reference | Reference |
| 2 | 1.0 (0.9, 1.2) | 1.0 (0.8, 1.2) | 1.1 (0.8, 1.3) | 1.0 (0.8, 1.2) | 1.0 (0.7, 1.2) | 1.0 (0.8, 1.3) |
| 3 | 1.3 (1.0, 1.6) | 0.9 (0.7, 1.3) | 1.3 (0.9, 1.7) | 1.2 (0.9, 1.5) | 0.8 (0.6, 1.1) | 1.2 (0.9, 1.6) |
| ≥4 | 0.9 (0.7, 1.1) | 0.8 (0.6, 1.2) | 0.9 (0.7, 1.2) | 0.8 (0.6, 1.0) | 0.7 (0.5, 1.1) | 0.8 (0.6, 1.1) |

Data are odds ratio (95%CI).
<sup>1</sup> Adjusted for sex, age, BMI, ethnicity, educational attainment, smoking status and number of comorbidity groupings (except when analysing this association).

Logistic regression models for potential COIVD-19 risk factors in COVID-19 +ve or -ve groups vs the background population group are presented in Table 4. After adjustment, both the COVID-19 +ve and -ve groups displayed greater odds of frailty, a number of frailty indicators and higher number of comorbidities, when compared with the background population group. The strength of associations were comparable in both groups.

**Table 4.**
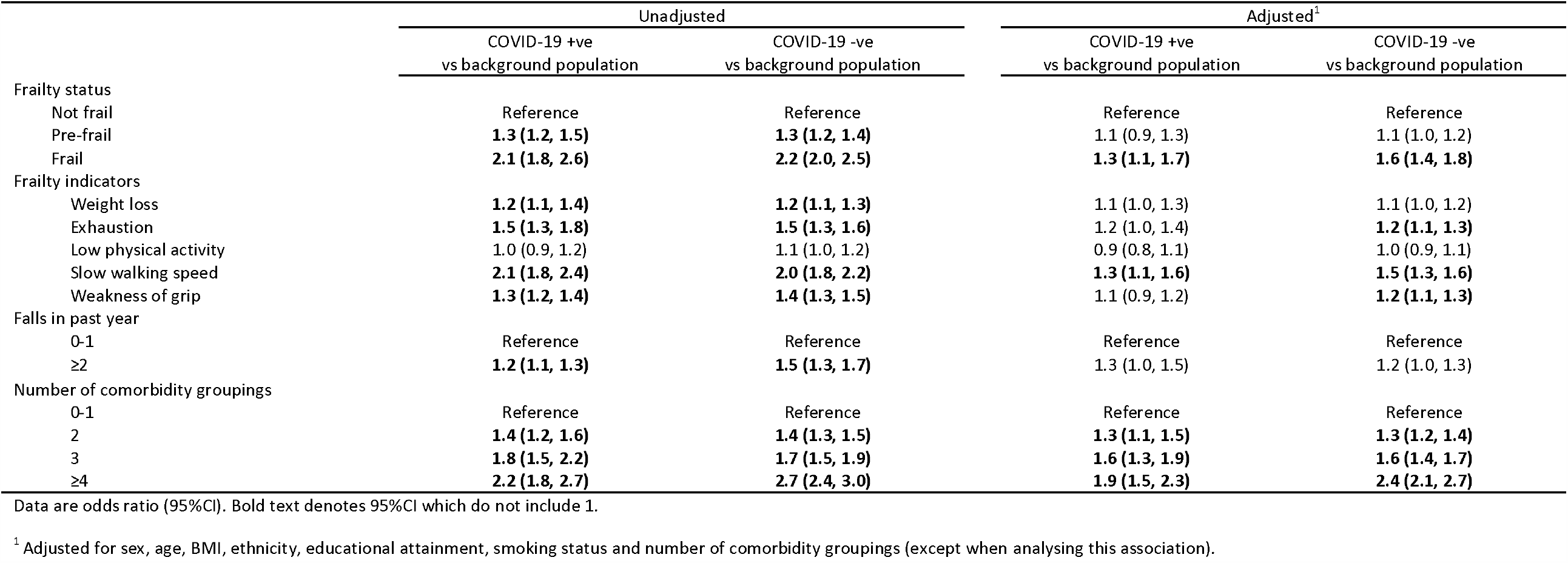
Associations for COVID-19 risk factors in COVID-19 +ve and COVID-19 -ve groups vs background population group.

## Discussion

To our knowledge and at the time of manuscript preparation, this is the first study to investigate associations between the frailty syndrome and COVID-19 diagnoses. Importantly, we have demonstrated that such classification may not aid risk stratification in terms of COVID-19 vulnerability, contrasting with other attributes, such as male sex, non-white ethnicity and greater BMI. Our findings also highlight that many characteristics of those hospitalised with COVID-19 disease are shared by those hospitalised for other reasons. However, it remains possible that factors such as frailty and multimorbidity may influence adverse outcomes following infection with SARS-CoV-2.

There are several limitations to this study. Firstly, the characteristics of participants used to calculate frailty and multimorbidity status for this study were recorded at recruitment into the UK Biobank, between 2006 and 2010. As such, participants may have accumulated markers of frailty or additional comorbidities after initial data collection, and so may be misclassified in the present analysis. The population studied was also relatively young, in terms of the wider frail population, when age was recorded at baseline [21]. However, given that such attributes develop over time, participants were substantially older (50-84 years) at time of COVID-19 testing, and we additionally analysed by age strata. Secondly, the COVID-19 test results used in this study, which are only from England, are also subject to limitations. Given that the majority of tests were undertaken in hospital, we cannot comment on the associations for asymptomatic or low severity COVID-19 cases within the community. Furthermore, the sensitivity of PCR-based testing has been reported as lower than chest CT imaging, potentially due to low viral load at the time of testing or inappropriate testing technique [22,23]. Therefore, the number of COVID-19 positive diagnoses within our sample may be underrepresented. Thirdly, at the time of manuscript preparation, mortality data for those tested for COVID-19 are not yet available within the UK Biobank resource. As such, we cannot comment on the associations for frailty and multimorbidity with COVID-19-associated mortality. Fourthly, records of clinical events occurring during hospital admissions are not available within the UK Biobank data set. Because of this, we also cannot comment on associations with adverse COVID-19 outcomes, such as non-invasive ventilation, intensive care admission or length of hospital stay. Finally, owing to the observational nature of this study causality cannot be inferred from our results.

Frailty is common, with global prevalence in those aged >85 years estimated to be 26% [21]. Frailty is characterised by a physiological vulnerability to stressor events, such as acute illnesses or hospital admissions, after which the individual fails to return to their previous baseline of health [24]. Ultimately, an individual living with frailty is predisposed to a significantly increased risk of hospital admission and higher rates of mortality [25]. Multimorbidity, defined as the presence of two or more chronic health conditions, is also common, with UK population estimates ranging from 15 to 30% [26]. Whilst multimorbidity is often present in those who are frail [27], it is also associated with greater risk of unplanned hospital admissions and increased mortality, independent of frailty [28,29]. Our results are in keeping with this existing literature regarding frailty and multimorbidity, with these populations being more susceptible to physiological insults such as COVID-19, and more likely to experience a severity of disease which requires hospitalisation. Importantly, our results suggest that people living with frailty and multimorbidity are no more likely to require a hospital admission due to COVID-19 compared to other conditions resulting in similar disease severity. Therefore, the reported high rates of COVID-19 diagnoses and mortality in those with multiple health conditions and those who are characterised as frail [4-7] are likely due to the highly contagious nature of COVID-19, and potential susceptibility to severe sequelae, rather than a specific propensity to contracting the disease.

The routine assessment of frailty during the COVID-19 pandemic has been frequently advocated, in order to facilitate appropriate management and resource allocation [8,30,10]. Additionally, in the UK, current NICE guidelines recommend the assessment of frailty as the initial step when assessing suspected COVID-19 patients on admission to hospital [11]. This assessment can then be used to inform decisions for escalation to intensive care environments where ventilatory support can be provided, in the case of patient deterioration. It has also been recommended that people living with multimorbidity or who are likely to be frail should minimise their exposure to the general population in order to reduce their risk of contracting COVID-19 [31]. To this end, Public Health England implemented a “shielding” strategy on 21^st^ March 2020, with particularly at risk patient groups being contacted based on underlying health conditions, and instructed to self-isolate until further notice [32]. We have not demonstrated any differences in frailty between those testing positive compared with those testing negative for COVID-19 (i.e. when comparing two groups presenting with disease, COVID-19 or otherwise, serious enough to warrant hospital admission). However, our findings do not allow comment on the predictive value of frailty for subsequent outcomes of COVID-19. They do, however, demonstrate that such individuals are generally at high risk of hospitalisation and requiring testing for COVID-19, and therefore risk minimisation for older frail or multimorbid individuals remains highly appropriate.

Finally, it is important not to view older age as synonymous with frailty when used as a potential risk factor for COVID-19. Whilst the majority of people living with frailty are older persons [21], a notable proportion of frail individuals are middle-aged [14]. The age range at baseline within our sample was 40-69 years, but it is important to note that this was during 2006-2010, and the age range at COVID-19 diagnosis was substantially older (50-84 years). Furthermore, although not statistically significant, the pattern of associations stratified by <60 or ≥60 years at baseline was consistent with the notion that frailty markers (here potentially assessed early in their development) might be more relevant in those contracting the COVID-19 at older ages.

## Conclusions

This is the first study to investigate associations between the frailty syndrome, multimorbidity and COVID-19 diagnoses within a large and well characterised prospective observational cohort. We have shown that no differences were evident for frailty status or number of morbidities when comparing those who tested positive for COVID-19 and those who tested negative, suggesting that the associations compared to the background population represent propensity to disease requiring hospital admission, rather than COVID-19 positivity per se. Studies are now urgently needed to examine the prognostic value of frailty and multimorbidity for adverse clinical sequelae following SARS-CoV-2 infection.

## Data Availability

Approval for access to data used in this study is via UK Biobank (approved Access Application 3593).

## Acknowledgments

This work was supported by the UK Medical Research Council, Wellcome Trust, National Institute for Health Research, Versus Arthritis, Royal Osteoporosis Society Osteoporosis and Bone Research Academy, International Osteoporosis Foundation, NIHR Southampton Biomedical Research Centre, University of Southampton, University Hospital Southampton NHS Foundation Trust and NIHR Oxford Biomedical Research Centre. EMC is supported by the Wellcome Trust (201268/Z/16/Z). SJW and SD are joint first author. CC and NCH are joint senior author.

## Declarations

### Funding (information that explains whether and by whom the research was supported)

This work was supported by the UK Medical Research Council, Wellcome Trust, National Institute for Health Research, Versus Arthritis, Royal Osteoporosis Society Osteoporosis and Bone Research Academy, International Osteoporosis Foundation, NIHR Southampton Biomedical Research Centre, University of Southampton, University Hospital Southampton NHS Foundation Trust and NIHR Oxford Biomedical Research Centre. EMC is supported by the Wellcome Trust (201268/Z/16/Z).

### Conflicts of interest/Competing interests (include appropriate disclosures)

The authors report no conflicting or competing interests in relation to this work.

### Ethics approval (include appropriate approvals or waivers)

This study had ethics approval as part of overall UK Biobank ethics approval (NHS National Research Ethics Service 16/NW/0274). We undertook the study under UK Biobank Access Application 3593.

### Consent to participate (include appropriate statements)

All UK Biobank participants gave written informed consent for data collection, analysis, and linkage.

### Consent for publication (include appropriate statements)

All UK Biobank participants gave written informed consent for data collection, analysis, and linkage.

### Code availability (software application or custom code)

All analyses were performed with Stata v 15.1 (StataCorp, College Station, Texas, USA). No custom code was used.

### Authors’ contributions (optional: please review the submission guidelines from the journal whether statements are mandatory)

SJW and SD performed the analysis and created the original manuscript, with senior supervision from NCH. CMP gave expert statistical advice. EMC, KW, EMD, HP and CC contributed epidemiological and clinical musculoskeletal/gerontological expertise. All authors reviewed and approved the final manuscript, providing comments and amendments.

---

**Supplementary Table 1.** Comorbidity groupings.

| Comorbidity grouping<sup>1</sup> | Conditions included |
| --- | --- |
| 1. Painful conditions | Back pain<br>Joint pain<br>Headaches (not migraine)<br>Sciatica<br>Plantar fasciitis<br>Carpal tunnel syndrome<br>Fibromyalgia<br>Arthritis<br>Shingles<br>Disc problem<br>Prolapsed disc/slipped disc<br>Spine arthritis/spondylitis<br>Ankylosing spondylitis<br>Back problem<br>Osteoarthritis<br>Gout<br>Cervical spondylosis<br>Trigeminal neuralgia<br>Disc degeneration<br>Trapped nerve/compressed nerve |
| 2. Hypertension | Hypertension<br>Essential hypertension |
| 3. Depression | Depression<br>Postnatal depression |
| 4. Asthma | Asthma |
| 5. Coronary Heart Disease | Heart attack/MI<br>Angina |
| 6. Treated dyspepsia | Gastro-oesophageal reflux (GORD)/gastric reflux<br>Oesophagitis /Barrett's oesophagus<br>Gastric stomach ulcers<br>Gastric erosions/gastritis<br>Duodenal ulcer<br>Dyspepsia/indigestion<br>Hiatus hernia<br>Helicobacter pylori |
| 7. Diabetes | Diabetic nephropathy<br>Diabetic neuropathy/ulcers<br>Diabetes<br>Type 1 diabetes<br>Type 2 diabetes<br>Diabetic eye disease |
| 8. Thyroid disorders | Thyroid problem (not cancer)<br>Hyperthyroidism/thyrotoxicosis<br>Hypothyroidism/myxoedema<br>Graves' disease<br>Thyroid goitre<br>Thyroiditis |
| 9. Rheumatoid arthritis, other inflammatory polyarthropathies, systemic connective tissue disorders and systemic autoimmune disorders | Myositis/myopathy<br>Systemic Lupus Erythematosus<br>Connective tissue disorder<br>Sjogren's syndrome/sicca syndrome<br>Dermatopolymyositis<br>Scleroderma/systemic sclerosis<br>Rheumatoid arthritis<br>Psoriatic arthropathy<br>Dermatomyositis<br>Polymyositis<br>Polymyalgia Rheumatica<br>Malabsorption/coeliac disease |
| 10. Chronic Obstructive Pulmonary Disease (COPD) | COPD/chronic obstructive airways disease<br>Emphysema/chronic bronchitis<br>Emphysema |
| 11. Anxiety, other neurotic, stress related and somatoform disorders | Anxiety/panic attacks<br>Nervous breakdown<br>Post-traumatic stress disorder<br>Obsessive compulsive disorder |
|  | Stress |
|  | Insomnia |
|  | Psychological/psychiatric problem |
| 12. Irritable bowel syndrome | Irritable bowel syndrome |
| 13. Alcohol problems | Alcohol dependency |
|  | Alcoholic liver disease/alcoholic cirrhosis |
| 14. Other psychoactive substance abuse | Opioid dependency |
|  | Other substance abuse/dependency |
| 15. Treated constipation | Constipation |
| 16. Stroke and Transient Ischaemic Attack (TIA) | Stroke |
|  | TIA |
|  | Subarachnoid haemorrhage |
|  | Brain haemorrhage |
|  | Ischaemic stroke |
| 17. Chronic kidney disease | Polycystic kidney |
|  | Diabetic nephropathy |
|  | Renal/kidney failure |
|  | Renal failure requiring dialysis |
|  | Renal failure not requiring dialysis |
|  | Kidney nephropathy |
|  | Immunoglobulin A (IgA) nephropathy |
| 18. Diverticular disease of intestine | Diverticular disease/diverticulitis |
| 19. Atrial fibrillation | Atrial fibrillation |
| 20. Peripheral vascular disease | Peripheral vascular disease |
|  | Leg claudication/intermittent claudication |
| 21. Heart failure | Cardiomyopathy |
|  | Hypertrophic cardiomyopathy |
|  | Heart failure/pulmonary oedema |
| 22. Prostate disorders | Prostate problem (not cancer) |
|  | Enlarged prostate |
|  | Benign prostatic hypertrophy |
| 23. Glaucoma | Glaucoma |
| 24. Epilepsy | Epilepsy |
| 25. Dementia | Dementia/Alzheimer/cognitive impairment |
| 26. Schizophrenia (and related non-organic psychosis) and bipolar disorder | Schizophrenia |
|  | Mania/bipolar disorder/manic depression |
| 27. Psoriasis or eczema | Eczema/dermatitis |
|  | Psoriasis |
| 28. Inflammatory bowel disease | Inflammatory bowel disease |
|  | Crohn's disease |
|  | Ulcerative colitis |
| 29. Migraine | Migraine |
| 30. Chronic sinusitis | Chronic sinusitis |
| 31. Anorexia or bulimia | Anorexia, bulimia/other eating disorder |
| 32. Bronchiectasis | Bronchiectasis |
| 33. Parkinson's disease | Parkinson's disease |
| 34. Multiple sclerosis | Multiple sclerosis |
| 35. Viral Hepatitis | Infective/viral hepatitis |
|  | Hepatitis B |
|  | Hepatitis C |
|  | Hepatitis D |
|  | Hepatitis E |
| 36. Chronic liver disease | Oesophageal varices |
|  | Non infective hepatitis |
|  | Liver failure/cirrhosis |
|  | Primary biliary cirrhosis |
| 37. Osteoporosis | Osteoporosis |
| 38. Chronic fatigue syndrome | Chronic fatigue syndrome |
| 39. Endometriosis | Endometriosis |
| 40. Meniere disease | Meniere disease |
| 41. Pernicious Anaemia | Pernicious anaemia |
| 42. Polycystic ovaries | Polycystic ovaries |
| 43. Cancer | Lifetime diagnosis |

<sup>1</sup> Self-reported lifetime diagnoses by a doctor, during face-to-face interviews at study recruitment (UK Biobank data field 20002), apart from cancer, which was reported during touchscreen questionnaires (UK Biobank data field 2453). Based on previous work by Barnett and colleagues [17] and Nicholl and colleagues [18].

